# Stress and Depression in the Workplace of Educators in the Philippines

**DOI:** 10.1101/2021.04.22.21254017

**Authors:** Caren Casama Orlanda-Ventayen, Randy Joy Magno Ventayen

## Abstract

Workplace Stress and depression should be treated properly in order to maintain productive teaching as the noblest profession. Unmanaged stress and depression could lead to a serious outcome that affects the happy environment in the classroom. Thus, managing stress and avoiding depression in the workplace is one of the important situations that a teacher should aim in order to succeed. This study aims to determine the level of workplace stress and depression of the educators in the Philippines by gathering their profile, workplace stress, and the state of depression. A correlation was investigated if there is a significant difference in the profile to the workplace stress and state of depression. Based on the result of the study, teachers sometimes experience stress in the workplace, while some experience depression at some point in time. It is recommended that teachers should understand how to practice stress management and avoiding depression.

## 1. Introduction and Background of the Study

Teaching is the noblest profession because it is the source of all professionals in the world. Without teachers, there will be no knowledge that was imparted and transferred. In order to continuously improved the educational system, healthy teachers are needed to maintain a happy environment in the classroom. Thus, managing stress in the workplace is one of the important situations that a teacher should aim in order to succeed. Academic Workload should also be manageable to maximize the productivity of the teachers.

A few numbers of the suicide of Teachers in the Philippines provides alarming results in the education sector in the Philippines (BusinessMirror, 2018). Based on several media reports, the Department of Education is mourning over the death of a teacher and say that they will look into it and it is non-work related. The Department also clarifies that the workload should not be blamed for the teacher’s suicide because there are other factors that may contribute (Mateo, 2018; Reyes, 2018). While The Teachers’ Dignity Coalition (TDC) met with DepEd officials to discuss concerns over the supposed workload, it cited that the heavy burden of paperwork is one among the reasons of the teacher who hanged herself in one case of teachers’ suicide in 2018. (Mateo, 2018).

While the education sector refuses to correlate workload with the suicides, they still emphasize that it is a wake-up call for public school teachers to learn how to manage work pressures that reacting to news circulating on social media that heavy paperwork had prompted one multi-grade teacher to commit suicide. The Department of Education urged to lighten teacher workloads (Hernando-Malipot, 2018). Due to the numerous reports, the secretary of the Department of Education said that they have already reduced the workload of teachers, which includes clerical and paper works. The secretary added that they are currently studying how to unload further teachers, based on the news report (Terrazola, 2018).

### 1.1 The Objective of the Study

This study aims to determine the sentiments of primary and secondary school by gathering the teacher’s profile, Workplace Stress, and the State of Depression. A correlation will be investigated if there is a significant difference in the profile to the workplace stress and state of depression. This study also aims to ask teachers all over the country on their workloads and perception about teacher’s depression.

### 1.2 Statement of the Problem

1. What is the profile of the respondents in terms of:
  a. Age
  b. Sex
  c. Employment Status
  d. Employment Type
  e. Teaching Level
  f. Level of Education
  g. Salary Range
  h. Length of Service
  i. Region
2. What is the level of workplace stress?
3. What is the level of Depression Assessment?
4. Is there a significant difference between the profile of the respondents and workplace stress?
5. Is there a significant difference between the profile of the respondents and the depression assessment?

## 2. Literature Review

Teachers are the ones who teach the students to be the best they can be; they should also be a role model for the student. In most cases, teachers are helping the students to avoid wrong thinking such as depression (Shilubane et al., 2015). One result of the study shows that teachers overwhelmingly agreed that they should have a role in suicide prevention (Hatton et al., 2017).

Teachers’ suicide is not new; there have been reported cases in the past such as the 3 Chinese teachers in Hong Kong in 1994, which were preceded by the suicides of several students. In the investigated study, teachers are said to have a greater workload in addressing the needs of more troubled students, which may increase their stress levels. (Leung, 1994).

Non-teaching career is not excluded from the historical problems of suicide. Several studies conducted that linking workload in depression and suicidal thoughts. One study shows a review of studies of stress and occupational difficulties experienced by veterinary surgeons. The results show that Occupational stressors included long working hours, heavy workload, poor work-life balance, difficult client relations are stressors that contribute to depression and suicidal thoughts. (Mendoza, 2019a, 2019b; Platt et al., 2012).

Despite the reported cases in the Philippines, the result of the study is different from the United States of America, where teachers have the lowest suicide rate of any profession. While Workers in farming, fishing, and forestry jobs had the highest rate: 84.5 per 100,000 workers, the suicide rate among people in education, training and library jobs was 7.5 per 100,000 workers (McIntosh et al., 2016; Tiesman et al., 2015). Comparing the workload in the United States teachers, predictably, U.S. teachers also spend more time teaching in the classroom than their international one study shows that U.S. teachers in grades 10 through 12 spent an average of 1,076 hours teaching students each year, while the global average is just 655 hours (Organisation for Economic Co-operation and Development, 2007, 2012, 2014). Based on the study, the working hours of teachers outside the Philippines are 3 to 5 hours a day. The report covers all 34 OECD member countries as well as ten partner countries (Moeny, 2014).

### 2.1 The workload of Educators in the Philippines

When we say Educators in the Philippines, we are referring to all with a teaching career that includes teachers, instructors, and professors. Elementary Teachers are teaching from Grade 1 to 8, Junior High School Teachers are teaching Grade 9 to 10, and Senior High School Teachers are teaching Grade 11 and 12 (Department of Education, 2009). Those teaching in the Vocational Programs and Higher Educational Institutions are called Instructors or Professors depending on the rank (Department of Budget Management, 2012). The regular workload for Teachers is 6 hours a day or 30 hours a week for a full-time teacher, which still excludes other functions as a teacher (Department of Education, 2009). While in State University and Colleges, a typical full-time Instructor or Professor is handling 18 hours teaching load per week with full time teaching time excluding designation, research, and extension function. SUC professors are de-loaded by corresponding other functions such as research, extension (Tarlac State University, 2008) and privilege in ongoing graduate education. There are some faculties in the State University that has a maximum of 3 to 6 hours a week teaching load because of full-time Graduate Schooling (Commission on Higher Education, 2016) due to the K to 12 Transition in the Philippines.

### 2.2. Synthesis of Literature Review

Based on the brief literature review, it can be seen that stress and depression in the workplace can be caused by a heavy academic workload. Despite this, there are other factors that affect the actions, stress and depression can be a triggering factor. In the United States, the teacher has the lowest suicidal rate. A workload of teachers in the United States is higher than other OECD countries which are around 3 to 5 hours a day, while in the Philippines the regular workload of a teacher is 6 to 8 hours a day for Elementary and Secondary teachers and 3 to 4 hours a day for State University Instructors or Professors excluding research and extension function.

## 3. Methodology

The target respondents of this study are all the educators in the Philippines. All teachers are invited to participate in the survey by answering an online form that was distributed in CHED K to 12 Program Scholars group, TESDA Teachers group, and DepEd Tambayan Facebook Group. Purposive and convenience sampling was used in order to gather the respondents.

Participants were requested to complete the survey by posting the link in the three Facebook groups. The researcher shortens the link using bit.ly. The survey questionnaire was floated using Google Forms, and extracted in CSV format for analysis.

### 3.1 Statistical Treatment Used

Frequency and Percentage were used in the primary objectives such as the profile of the respondents. Average weighted mean was also used in determining the interpretation based on the Likert rating scale used.

For the last problem, A Pearson correlation was also used and measured Correlation that is significant at the 0.05 level (2-tailed). In order to simplify statistical computation, all data was inputted into the software SPSS for faster analysis of data.

## 4. The result of the Study

The result part of the study determines the sentiments of primary and secondary school by gathering the teacher’s profile, Workplace Stress, and the State of Depression. A Correlation will be investigated if there is a significant difference in the profile to the workplace stress and state of depression.

### Profile of Respondents

The majority of the respondents are female with 74 percent of the total respondents. This implies that there are more female educators in the Philippines compared to male educators.

As shown in the table, the majority of the respondents with a 92.1 percent have the security of tenure.

As shown in Table 4, the Majority of the respondents are working in the government sector.

**Table 1.** Likert Scale Used

| **Range** | **Descriptive Equivalent** | **Descriptive Equivalent** |
| --- | --- | --- |
| 3.25 to 4.00 | Strongly Agree | Often |
| 2.50 to 3.24 | Agree | Sometimes |
| 1.75 to 2.49 | Disagree | Seldom |
| 1.00 to 1.74 | Strongly Disagree | Never |

**Table 2.** Frequency and Percentage distribution of respondents in terms of sex

| **Sex** | **Frequency** | **Percentage** |
| --- | --- | --- |
| Male | 106 | 26 |
| Female | 301 | 74 |
| **TOTAL** | **407** | **100** |

**Table 3.** Frequency and Percentage Distribution of Respondents according to Employment Status

| **Employment Status** | **Frequency** | **Percentage** |
| --- | --- | --- |
| Regular/Permanent | 375 | 92.1 |
| Probationary/Temporary | 13 | 3.2 |
| Contractual/Contract of Service/Part-Time | 19 | 4.7 |
| **TOTAL** | **407** | **100** |

**Table 4.** Frequency and Percentage Distribution of Respondents in terms of Length Employment Type

| **Employment Type** | **Frequency** | **Percentage** |
| --- | --- | --- |
| Government | 369 | 90.7 |
| Private | 38 | 9.3 |
| **TOTAL** | **407** | **100** |

The teaching level of educators in the Philippines is distributed from the kindergarten level to the tertiary level.

As shown in Table 6, the majority of the respondents have ongoing Master’s units.

**Table 5.** Frequency and Percentage Distribution of Respondents in terms of Teaching Level

| **Teaching Level** | **Frequency** | **Percentage** |
| --- | --- | --- |
| Kindergarten | 6 | 1.5 |
| Elementary (Grade 1 to 3) | 55 | 13.5 |
| Elementary (Grade 4 to 8) | 96 | 23.6 |
| Junior High School (Grade 9 and 10) | 89 | 21.9 |
| Senior High School (Grade 11 and 12) | 77 | 18.9 |
| Vocational Level | 45 | 11.1 |
| Undergraduate / College | 39 | 9.6 |
| **TOTAL** | **407** | **100** |

**Table 6.** Frequency and Percentage Distribution of Respondents in terms of Level of Education

| **Level of Education** | **Frequency** | **Percentage** |
| --- | --- | --- |
| Vocational or Bachelor's Level | 1 | 2 |
| Bachelor's Graduate | 102 | 25.1 |
| Master Level | 150 | 36.9 |
| Masteral Graduate | 69 | 17 |
| Doctoral Level | 71 | 17 |
| Doctoral Graduate | 14 | 3.4 |
| **TOTAL** | **407** | **100** |

Table 7 shows the salary range of Teachers in the Philippines, where the majority of the respondents have 10,000 to 20,000, followed by 20,000 to 30,000.

**Table 7.** Frequency and Percentage Distribution of Respondents in terms of Salary Range

| **Salary Range** | **Frequency** | **Percentage** |
| --- | --- | --- |
| Less than 10,000 | 53 | 13 |
| 10,001 to 20,000 | 271 | 66.6 |
| 20,001 to 30,000 | 37 | 9.1 |
| 30,001 to 40,000 | 18 | 4.4 |
| 40,001 to 50,000 | 6 | 1.5 |
| 50,001 to 75,000 | 16 | 3.9 |
| 75,001 to 100,000 | 4 | 1.0 |
| **TOTAL** | **405** | **99.5** |

**Table 8.** Frequency and Percentage Distribution of Respondents in terms of Preference of Length of Service

| **Length of Service** | **Frequency** | **Percentage** |
| --- | --- | --- |
| Less than a year | 24 | 5.9 |
| 1 to 2 years | 41 | 10.1 |
| 2 to 5 years | 107 | 26.4 |
| 6 to 10 years | 104 | 25.6 |
| 11 to 15 years | 67 | 16.5 |
| 16 to 20 years | 25 | 6.2 |
| 21 to 30 years | 32 | 7.9 |
| 30 years and above | 6 | 1.5 |
| **TOTAL** | **406** | **100** |

**Table 9.** Frequency and Percentage Distribution of Respondents in terms of Region

| **Highest Academic** | **Frequency** | **Percentage** |
| --- | --- | --- |

| **Qualification** |  |  |
| --- | --- | --- |
| Region 1 | 71 | 17.4 |
| CAR | 13 | 3.2 |
| NCR | 33 | 8.1 |
| Region 2 | 18 | 4.4 |
| Region 3 | 38 | 9.3 |
| Region 4A | 67 | 16.5 |
| Region 4B | 6 | 1.5 |
| Region 5 | 9 | 2.2 |
| Region 6 | 21 | 5.2 |
| Region 7 | 56 | 13.8 |
| Region 8 | 41 | 10.1 |
| Region 9 | 27 | 6.6 |
| CARAGA | 4 | 1.0 |
| ARMM | 1 | 0.2 |
| **TOTAL** | **406** | **100** |

**Table 10.**
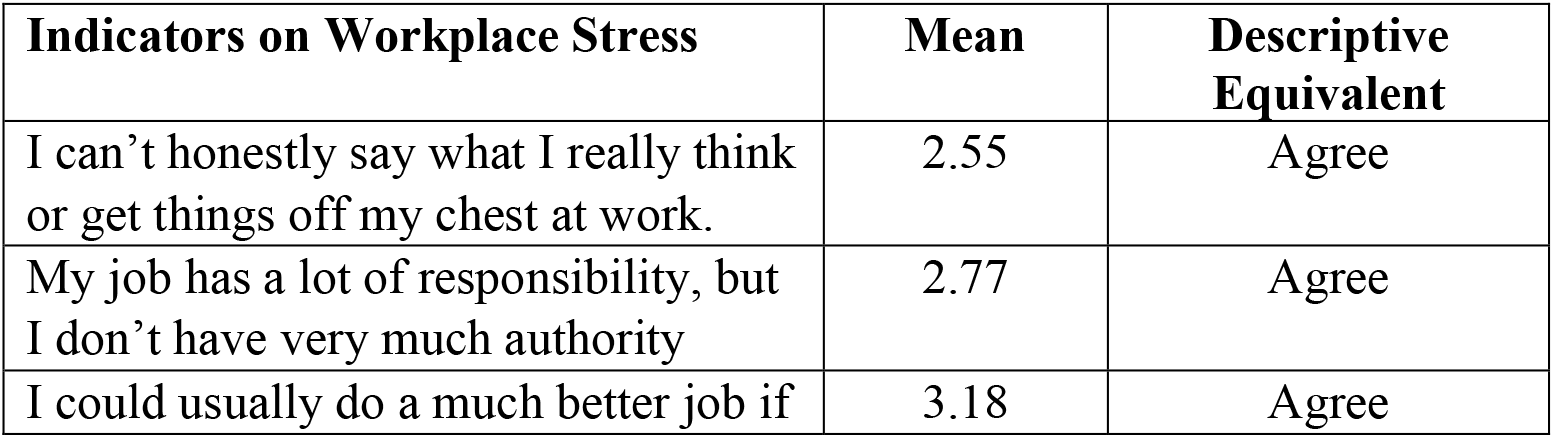
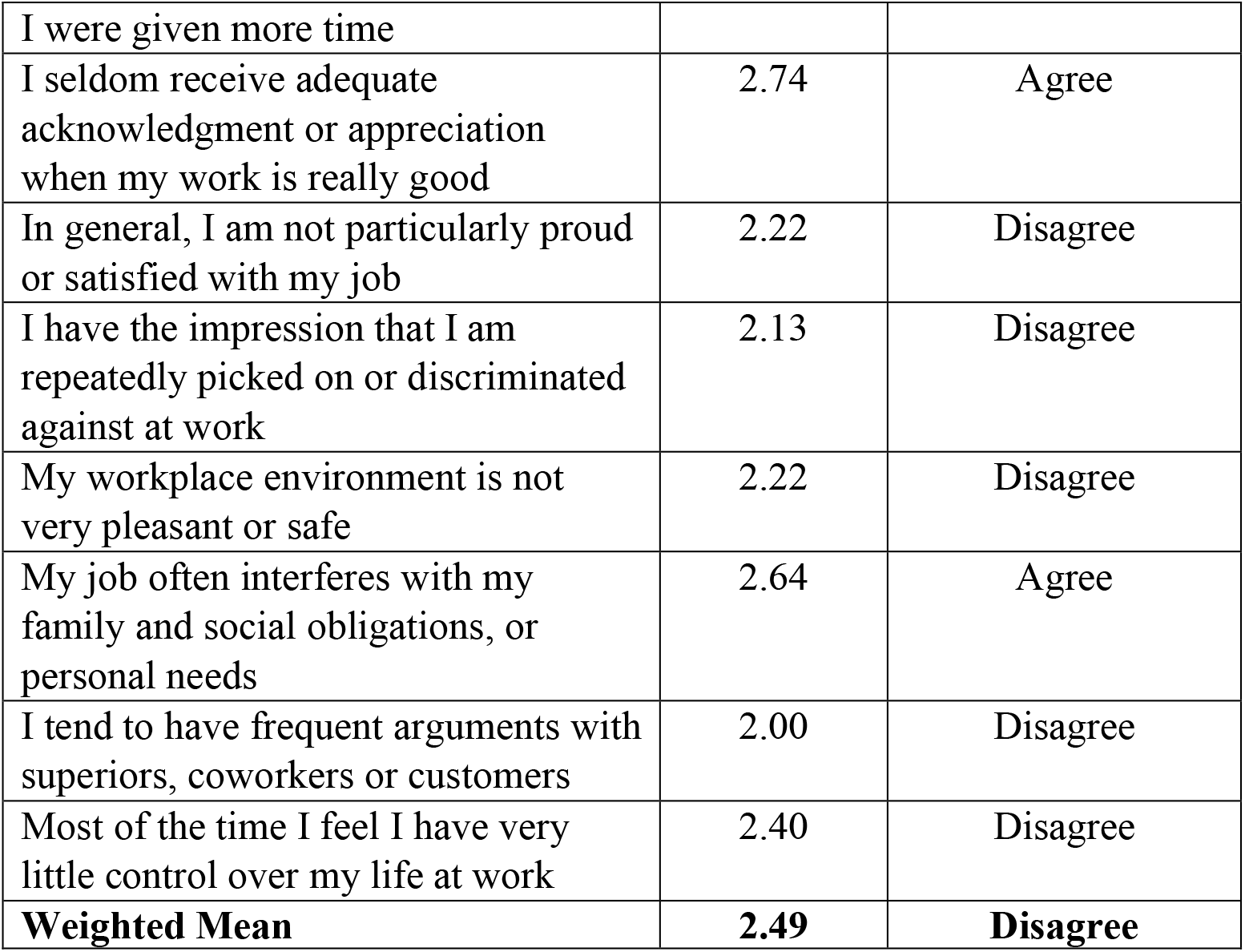
Level of Workplace Stress of Teachers

The majority of the respondents are engaged in teaching from 2 to 5 years.

The distribution of respondents shows that all regions are distributed.

The workplace stress of teachers is shown in the table, where most of the teachers need more time in order to finish the required task given to them. As stated in the literature review, the regular workload for teachers in elementary and secondary level is 6 hours a day or 30 hours a week, which still excludes other functions as a teacher (Department of Education, 2009). This implies that the workload of educators in the Philippines is overloaded and teachers need more time to finish the task assigned to them. Most of the educators agree that work is continuous as a teacher and does not practice their authority over their work where lack of acknowledgment and appreciation is also an issue as there is an impact of autocratic and democratic leadership style on their job satisfaction (Bhatti et al., 2012).

Workload also interferes with the family and social obligations of the teachers, where some of the teachers are bringing their paperwork at home. 75 percent of school hours are consumed in teaching and the remaining 25 percent of school hours are dedicated to paper works.

Workplace depression of teachers is shown in table 11, where teachers are bothered by feeling tired or having little energy over the last two weeks. It is also shown that teachers do not have a severe depression towards work since most of them are educated and may be able to handle the situation even in hard times.

**Table 11.**
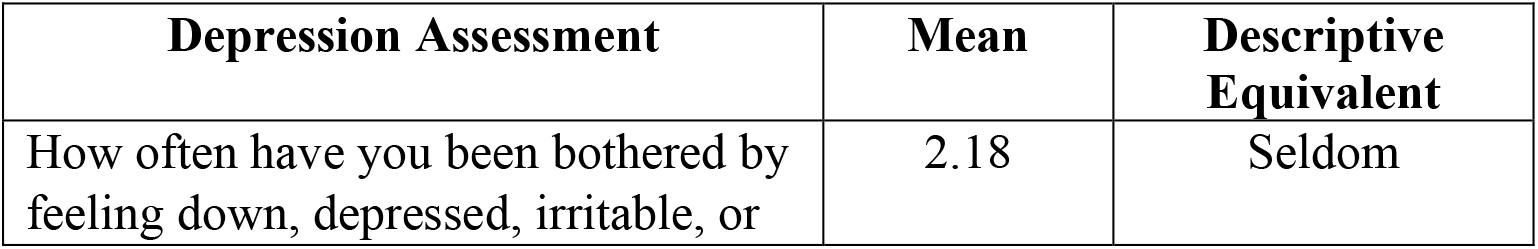
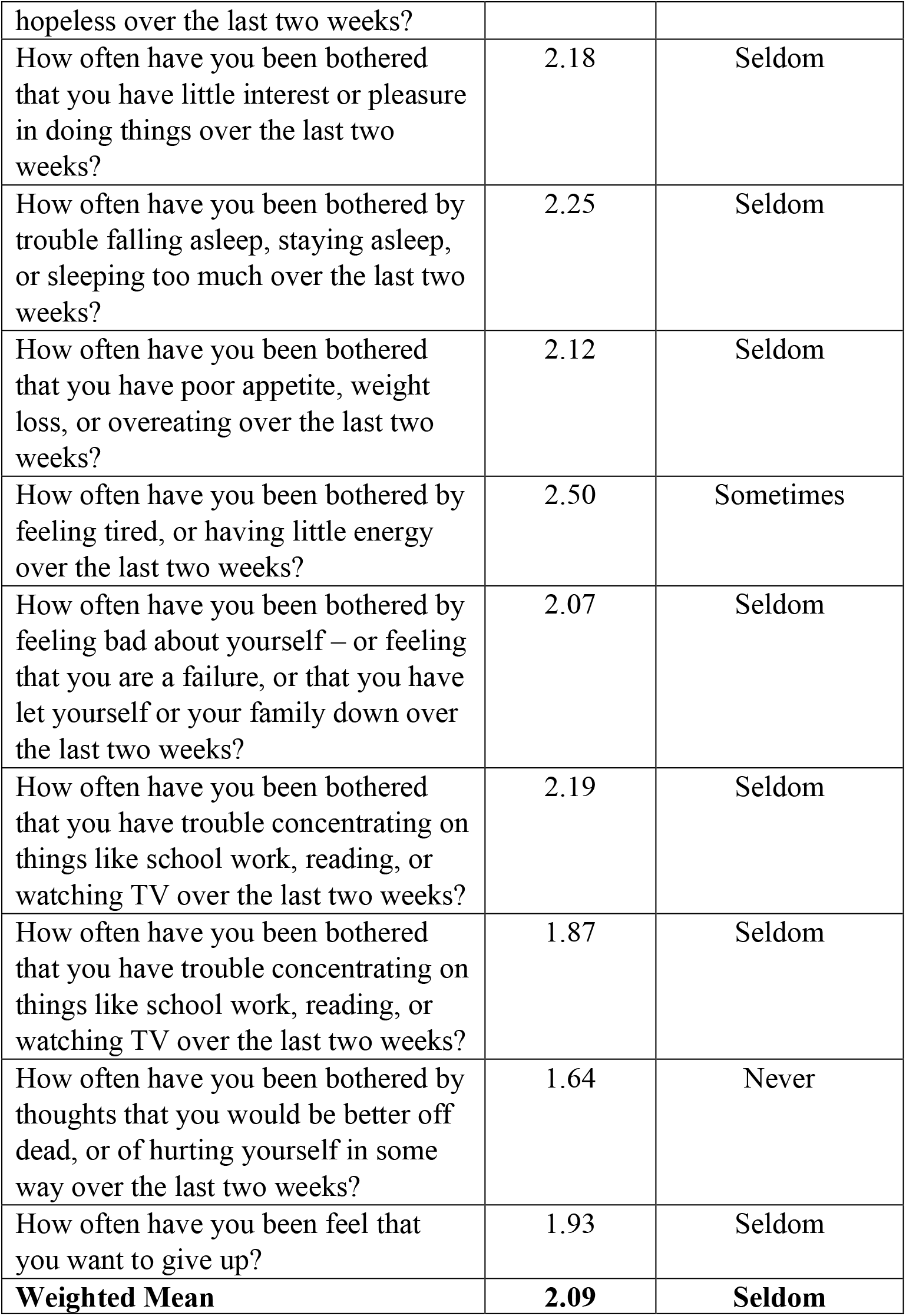
Level of Depression of the Teachers

**Table 12.** Significant Difference between Level of Workplace Stress and profile variables

| Profile Variables | Groups | Mean | Result | F-value | Sig. Level | Interpretation |
| --- | --- | --- | --- | --- | --- | --- |
| Sex | Male | 2.42 | Disagree | 1.187 | 0.277 | Not Significant |
|  | Female | 2.51 | Agree |  |  |  |
| Employment Status | Regular/Permanent | 2.49 | Disagree | 0.118 | 0.889 | Not Significant |
|  | Probationary/ Temporary | 2.40 | Disagree |  |  |  |
|  | Contractual/ Contract of Service/Part-Time | 2.48 | Disagree |  |  |  |
| Employment Type | Government | 2.50 | Agree | 3.210 | 0.074 | Not Significant |
|  | Private | 2.30 | Disagree |  |  |  |
| Teaching Level | Kindergarten | 2.85 | Agree | 5.016 | 0.00 | Significant |
|  | Elementary (Grade 1 to 3) | 2.46 | Disagree |  |  |  |
|  | Elementary (Grade 4 to 8) | 2.49 | Disagree |  |  |  |
|  | Junior High School (Grade 9 and 10) | 2.63 | Agree |  |  |  |
|  | Senior High School (Grade 11 and 12) | 2.62 | Agree |  |  |  |
|  | Vocational Level | 2.22 | Disagree |  |  |  |
|  | Undergraduate / College | 2.14 | Disagree |  |  |  |
| Level of Education | Vocational or Bachelor's Level | 2.60 | Agree | 3.185 | 0.008 | Significant |
|  | Bachelor's Graduate | 2.62 | Agree |  |  |  |
|  | Masteral Level | 2.55 | Agree |  |  |  |
|  | Masteral Graduate | 2.33 | Disagree |  |  |  |
|  | Doctoral Level | 2.31 | Disagree |  |  |  |
|  | Doctoral Graduate | 2.34 | Disagree |  |  |  |
| Salary Range | Less than 10,000 | 2.50 | Agree | 2.192 | 0.043 | Significant |
|  | 10,001 to 20,000 | 2.52 | Agree |  |  |  |
|  | 20,001 to 30,000 | 2.25 | Disagree |  |  |  |
|  | 30,001 to 40,000 | 2.13 | Disagree |  |  |  |
|  | 40,001 to 50,000 | 2.05 | Disagree |  |  |  |
|  | 50,001 to 75,000 | 2.61 | Agree |  |  |  |
|  | 75,001 to 100,000 | 2.98 | Agree |  |  |  |
| Length of Service | Less than a year | 2.38 | Disagree | 1.197 | 0.303 | Not Significant |
|  | 1 to 2 years | 2.53 | Agree |  |  |  |
|  | 2 to 5 years | 2.47 | Disagree |  |  |  |
|  | 6 to 10 years | 2.57 | Agree |  |  |  |
|  | 11 to 15 years | 2.55 | Agree |  |  |  |
|  | 16 to 20 years | 2.38 | Disagree |  |  |  |
|  | 21 to 30 years | 2.32 | Disagree |  |  |  |
|  | 30 years and above | 2.05 | Disagree |  |  |  |
| Region | Region 1 | 2.40 | Disagree | 0.632 | 0.827 | Not Significant |
|  | CAR | 2.49 | Disagree |  |  |  |
|  | NCR | 2.63 | Agree |  |  |  |
|  | Region 2 | 2.56 | Agree |  |  |  |
|  | Region 3 | 2.52 | Agree |  |  |  |
|  | Region 4A | 2.44 | Disagree |  |  |  |
|  | Region 4B | 2.13 | Disagree |  |  |  |
|  | Region 5 | 2.21 | Disagree |  |  |  |
|  | Region 6 | 2.52 | Agree |  |  |  |
|  | Region 7 | 2.49 | Disagree |  |  |  |
|  | Region 8 | 2.54 | Agree |  |  |  |
|  | Region 9 | 2.62 | Agree |  |  |  |
|  | CARAGA | 2.38 | Disagree |  |  |  |
|  | ARMM | 2.70 | Agree |  |  |  |

**Table 13.** Significant Difference between Frequency of Depression and profile variables

| **Profile Variables** | **Groups** | **Mean** |  | **F-value** | **Sig. Level** | **Interpretation** |
| --- | --- | --- | --- | --- | --- | --- |
| Sex | Male | 2.09 | Seldom | 0.011 | 0.915 | Not Significant |
|  | Female | 2.10 | Seldom |  |  |  |
| Employment Status | Regular/Permanent | 2.08 | Seldom | 1.57 | 0.21 | Not Significant |
|  | Probationary/Temporary | 2.08 | Seldom |  |  |  |
|  | Contractual/Contract of Service/Part-Time | 2.40 | Seldom |  |  |  |
| Employment Type | Government | 2.17 | Seldom | 4.46 | 0.035 | Significant |
|  | Private | 1.84 | Seldom |  |  |  |
| Teaching Level | Kindergarden | 2.17 | Seldom | 6.44 | 0.000 | Significant |
|  | Elementary (Grade 1 to 3) | 2.27 | Seldom |  |  |  |
|  | Elementary (Grade 4 to 8) | 2.22 | Seldom |  |  |  |
|  | Junior High School (Grade 9 and 10) | 2.33 | Seldom |  |  |  |
|  | Senior High School (Grade 11 and 12) | 2.05 | Seldom |  |  |  |
|  | Vocational Level | 1.72 | Never |  |  |  |
|  | Undergraduate / College | 1.61 | Never |  |  |  |
| Level of Education | Vocational or Bachelor's Level | 2.60 | Sometimes | 7.90 | 0.000 | Significant |
|  | Bachelor's Graduate | 2.62 | Sometimes |  |  |  |
|  | Masteral Level | 2.55 | Sometimes |  |  |  |
|  | Masteral Graduate | 2.33 | Seldom |  |  |  |
|  | Doctoral Level | 2.31 | Seldom |  |  |  |
|  | Doctoral Graduate | 2.34 | Seldom |  |  |  |
| Salary Range | Less than 10,000 | 2.09 | Seldom | 4.29 | 0.000 | Significant |
|  | 10,001 to 20,000 | 2.14 | Seldom |  |  |  |
|  | 20,001 to 30,000 | 1.83 | Seldom |  |  |  |
|  | 30,001 to 40,000 | 1.66 | Never |  |  |  |
|  | 40,001 to 50,000 | 1.31 | Never |  |  |  |
|  | 50,001 to 75,000 | 2.52 | Sometimes |  |  |  |
|  | 75,001 to 100,000 | 2.78 | Sometimes |  |  |  |
| Length of Service | Less than a year | 2.25 | Seldom | 1.28 | 0.257 | Not Significant |
|  | 1 to 2 years | 2.25 | Seldom |  |  |  |
|  | 2 to 5 years | 2.14 | Seldom |  |  |  |
|  | 6 to 10 years | 2.08 | Seldom |  |  |  |
|  | 11 to 15 years | 2.09 | Seldom |  |  |  |
|  | 16 to 20 years | 1.74 | Never |  |  |  |
|  | 21 to 30 years | 2.0 | Seldom |  |  |  |
|  | 30 years and above | 1.95 | Seldom |  |  |  |
| Region | Region 1 | 2.23 | Seldom | 0.951 | 0.50 | Not Significant |
|  | CAR | 2.24 | Seldom |  |  |  |
|  | NCR | 2.17 | Seldom |  |  |  |
|  | Region 2 | 2.09 | Seldom |  |  |  |
|  | Region 3 | 2.13 | Seldom |  |  |  |
|  | Region 4A | 1.90 | Seldom |  |  |  |
|  | Region 4B | 1.95 | Seldom |  |  |  |
|  | Region 5 | 1.61 | Never |  |  |  |
|  | Region 6 | 2.01 | Seldom |  |  |  |
|  | Region 7 | 2.18 | Seldom |  |  |  |
|  | Region 8 | 2.11 | Seldom |  |  |  |
|  | Region 9 | 1.99 | Seldom |  |  |  |
|  | CARAGA | 2.22 | Seldom |  |  |  |
|  | ARMM | 2.30 | Seldom |  |  |  |

A number of suicide of educators is alarming results in the education sector of the Philippines (BusinessMirror, 2018). The Department of Education clarifies that the workload should not be blamed for the teacher’s suicide because there are other factors that may contribute (Mateo, 2018; Reyes, 2018). While The Teachers’ Dignity Coalition (TDC) met with DepEd officials to discuss concerns over the supposed workload, the department was urged to lighten teacher workloads (Hernando-Malipot, 2018). The result of the study does not support that workload should be blamed in the depression of the teachers who committed suicides. There is no direct effect of the workload, though it may be a contributory factor that a teacher might end his or her life due to workload.

The table shows that there is a significant difference between the level of workplace stress across teaching level, educational attainment and salary range.

Based on the result of the study, the level of stress is high for the teachers teaching in Kindergarten, followed by the Junior and Senior levels compared to the elementary grade. It was also shown that Workplace stress is lower in vocational and college level compared to any other educational level. A typical full-time Instructor or Professor is handling 18 hours and are de-loaded by corresponding other functions such as research, extension (Tarlac State University, 2008), while the teacher in the kindergarten up to the senior high school are teaching 30 hours per week with an average of 6 hours a day.

Bachelor’s graduate and master’s level teachers have a higher level of workplace stress compared to those who are graduate with a master’s degree and doctoral degree. Likewise, those earning below 20,000 has a higher level of stress compared to those who are earning 20,000 and above. The result of the study agrees with several studies that salary has a contribution to the level of stress of workers (Muhammad Shahzad Chaudhry; Hazoor Muhammad Sabir; Rafi, 2011; Parvin & Karbin, 2011; Teck-Hong & Waheed, 2011).

The table shows that there is a significant difference between the level of workplace depression across employment type, teaching level, educational attainment, and salary range.

Based on the result of the study, the level of depression is high for the teachers teaching in the government sector than in the private institutions. It was also presented that Junior and elementary grade levels are more depressed than other educators. It was also shown that Workplace depression is lower in vocational and college level compared to any other educational level. A workload and stress may have a contributory factor in the level of depression especially among those working full time (Caspi et al., 2003; Kubo, 2007; Van Praag, 2004).

Bachelor’s graduate and master’s level teachers have a higher level of depression compared to those who are graduate in a master’s degree and doctoral degree. Likewise, those earning below 20,000 has a higher level of depression compared to those who are earning 20,000 and above. Despite the result of the study shows that those earning 50,000 and above have a higher level of depression compared to the other range, the limitation of this research is very limited to the few numbers of respondents that may not represent the overall population.

Based on the overall result of the study, stress and depression of teachers are connected to each other, but as a limitation of this study, causes are not identified. Stress is the body’s response to physical or emotional demands and emotional stress can play a role in causing depression or be a symptom of it. A stressful situation, especially in the workload of the teachers, can trigger feelings of depression, and these feelings can make it more difficult to deal with stress.

## Conclusions and Recommendations

Teaching is the noblest profession because it is the source of all professionals in the world. Without teachers, there will be no knowledge that was imparted and transferred. As a conclusion of this study, the level of stress and depression is high for the teachers teaching at lower levels such as kindergarten, elementary and high school. It was also shown that Workplace stress and depression is lower in vocational and college level. It is recommended that the government and administrator should look at the possible intervention to minimize the level of stress and depression of educators.

## Data Availability

no other data available in the internet

